# Efficacy and Mechanisms of Cannabis Oil for Alleviating Side Effects of Breast Cancer Chemotherapy (CBC2): Protocol for Randomized Controlled Trial

**DOI:** 10.1101/2023.01.01.23284097

**Authors:** May Soe Thu, Krit Pongpirul, Mawin Vongsaisuwon, Chanida Vinayanuwattikun, Kamonwan Banchuen, Thunnicha Ondee, Sunchai Payungporn, Phanupong Phutrakool, Preecha Nootim, Pajaree Chariyavilaskul, Sarocha Cherdchom, Kulthanit Wanaratna, Nattiya Hirankarn

## Abstract

**Background:** In a pilot study using both cannabidiol (CBD) and tetrahydrocannabinol (THC) as single agents in advanced cancer patients undergoing palliative care in Thailand, the doses were generally well tolerated, and the outcome measure of total symptom distress scores showed overall symptom benefit. The current study aims to determine the intensity of the symptoms experienced by breast cancer patients receiving chemotherapy, to explore the microbiome profile, cytokines, and bacterial metabolites before and after the treatment with cannabis oil or no cannabis oil, and to study the pharmacokinetics parameters and pharmacogenetics profile of the doses.

**Methods:** A randomized, double-blinded, placebo-controlled trial will be conducted on the metastatic breast cancer cases receiving chemotherapy at King Chulalongkorn Memorial Hospital (KCMH), Bangkok, Thailand. Block randomization will be used to allocate the patients into three groups: Ganja Oil (THC 2 mg/ml; THC 0.08 mg/drop, and CBD 0.02 mg/drop), Metta Osot (THC 81 mg/ml; THC 3 mg/drop), and placebo oil. The Edmonton Symptom Assessment System (ESAS), microbiome profile, cytokines, and bacterial metabolites will be assessed before and after the interventions.

**Thai Clinical Trial Registration:** TCTR20220809001

## Introduction

Breast cancer patients reported lower quality of life (QoL) globally (1, 2, 3), and chemotherapy was a factor in worsening QoL among the patients (4). Despite standard medical treatment, the impact of integrating complementary and alternative medicine was introduced into the current practice (5). Therefore, cannabinoids from *Cannabis sativa* have become attractive due to their major impact in reducing pain associated with chemotherapy and tumor-associated symptoms (6, 7, 8).

In a pilot study using both cannabidiol (CBD) and tetrahydrocannabinol (THC) as single agents in advanced cancer patients undergoing palliative care, the doses of THC and CBD were generally well tolerated, and the outcome measure of total symptom distress scores (TSDSs) showed overall symptom benefit (9). The average daily amounts tolerated were 300 mg of CBD and 10 mg of THC, respectively (range 5–30 mg), and 43% met the response criteria of ≥6 point reduction in TSDSs (9).

Several clinically relevant mouse models of breast cancer have shown that naturally sourced cannabinoids possess anti-tumor activities, with CBD as the most potent inhibitor of cancer cell proliferation (10). In experimental models, psychoactive THC and non-psychoactive CBD disturbed the disease progression and its effects on signaling pathways within cancer cells following activation of G-protein coupled cannabinoid-receptors (CB-Rs), other receptors such as GPR55 and TRPV1, and in a receptor-independent way. Both *in vitro* and *in vivo* studies have revealed that cannabinoids can activate these receptors, leading to anti-tumorigenic activity in most cases (11).

The human microbiota has recently raised so much attention as a significant risk modulator since they play a prominent role in regulating steroid hormone metabolism through stimulating different enzymes, such as hydroxysteroid dehydrogenase (12, 13). Despite the lack of convincing evidence that breast cancer is caused by dysbiosis, the comparison of several breast samples reveals variations in the abundance and microbial diversity of some particular genera between healthy individuals and patients (14, 15). Additionally, it recognizes that various breast cancer subtypes are correlated to multiple intestinal microbial signatures (16).

It is linked to both the dysbiosis of the intestinal microbiota and the intestinal dysfunction that results in inflammatory and immunological ailments. This implies that diseases spread through microbial translocation. It was proposed that the gut microbiota spreads to the skin before making its way to the breast and that dendritic cells may carry the bacteria there (17). Therefore, exploring the bacterial translocation markers to understand this unclear underlying mechanism becomes critical.

Additionally, less fungal diversity and richness has been seen in cancer patients (18), but it hasn’t been thoroughly investigated if particular species and their particular metabolic outputs characterize and support carcinogenesis. varied hormonal subtypes of breast cancer exhibit varied fungal signatures in the tumor microenvironment, and individuals with estrogen receptor-positive breast cancer showed higher levels of fungal variety than those with triple-negative breast cancer (16, 19).

Because of the primary interactions between skin microorganisms and the underlying mucosal immune function, the function of the breast skin microbiota is still unknown despite recent developments. Metastases and breast cancer have been linked to high abundances of skin commensals, particularly multiple species of *Staphylococcus*, in a thorough investigation of the skin microbiota in breast cancer (20). Several pathways have been hypothesized for microbial transfer to underlying tissue, including retrograde transfer through ductal networks, permeabilization of the epidermal barrier, and migration through the nipple-aspirate fluid (20).

There are hints that breast cancer and the oral microbiome are related. Women with periodontal disease, which is brought on by particular bacteria like the red complex (*Porphyromonas gingivalis, Tannerella forsythia, and Treponema denticola*), and the orange complex (*Fusobacterium nucleatum, Prevotella intermedia, Prevotella nigrescens, Peptostreptococcus micros, Streptococcus constellatus, Eubacterium nodatum, Campylobacter showae, Campylobacter gracilis, and Campylobacter rectus*), have been found to have an increased risk of breast cancer in some studies (21, 22, 23).

Currently, the Herb and Thai Traditional Medicine Development Division, Department of Thai Traditional and Alternative Medicine (DTAM), Ministry of Public Health, Thailand, manufactures Metta Osot (THC 81 mg/ml; THC 3 mg/drop) and Ganja Oil (THC 2 mg/ml; THC 0.08 mg/drop and CBD 0.02 mg/drop), both of which are produced under license number 13/2562. The preliminary analysis of the Thai Cannabis Practice Patterns and QoL (Thai Cannabis PQ) study assessed the pain symptoms using the Edmonton Symptom Assessment System (ESAS) and QoL using the EuroQoL Group’s 5-dimension, 5-level (EQ-5D-5L). It revealed that cancer patients had significantly lower baseline EQ-5D-5L (0.80±0.29 vs. 0.85±0.22) and ESAS pain (3.44±3.21 vs. 4.05±3.30) and anxiety (2.03±2.77 vs. 2.37±2.85) but higher tiredness (3.25±3.08 vs. 2.89±2.83), nausea (1.00±2.07 vs. 0.69±1.66), poor appetite (2.21±2.94 vs. 1.20±2.20), and shortness of breath (1.74±2.61 vs. 1.22±2.17) than non-cancer individuals (24). Ganja Oil and Metta Osot were added to the National List of Essential Medicines (NLEM) (25).

The pharmacokinetic profile of cannabinoids depends on routes of medication such as oral dosage and inhalation. A study revealed that the median value of the time taken to achieve the highest levels (T_max_) of sublingual drops for CBD only and a combination of CBD and THC was 2.17 hours (range 1 – 4 hours) and 1.67 hours (range 1 – 3 hours) and the mean of the highest concentration of the drops (C_max_) was 2.05 (SD = 0.92) and 2.58 (SD = 0.68) nanograms/ml, respectively (26).

Hence, the current randomized study aims to explore the efficacy of Thai cannabis oil products on the QoL of individuals diagnosed with advanced breast cancer and treated with chemotherapies and the adverse effects of Thai cannabis oil, along with the study of microbial alteration and diversity by cannabis among those patients. We aim to determine the intensity of the pain and QoL experienced by cancer patients using the ESAS and the EQ-5D-5L forms, respectively, to explore the microbiome profile, cytokines, and bacterial metabolites in hormone-dependent and independent breast cancer patients at baseline, to study the alteration of microbiome profile, cytokines, and bacterial metabolites before and after the treatment with the cannabis oil or no cannabis, and to study the pharmacokinetics parameters and pharmacogenetics profile.

## Materials and methods

### Target population

We include 90 adult breast cancer women patients currently receiving chemotherapy at King Chulalongkorn Memorial Hospital (KCMH) with an anticipated starting recruitment approximately in June 2023. The sample size was calculated using the mean ESAS values of the previous study (24).

#### Inclusion criteria

1. Women aged 18-70 years old who were diagnosed with metastatic breast cancer
2. Metastatic breast cancer patients who are currently receiving different doses of chemotherapy regimen with high/low emetic risk such as AC (cyclophosphamide and doxorubicin), AC followed by T (cyclophosphamide, doxorubicin, and paclitaxel), DC (cyclophosphamide and docetaxel), CMF (cyclophosphamide, fluorouracil, and methotrexate), and Paclitaxel
3. Breast cancer patients at KCMH during the study period
4. Participants provided written informed consent
5. Individuals having at least each score of ESAS parameters ≥3

#### Exclusion criteria

1. Women who have contraindications for chemotherapy, such as anaphylaxis
2. Women who have serious chemotherapy complications such as anaphylactic shock, bone marrow suppression, liver failure, acute kidney injury, and cardiac arrest
3. Women who are scheduled for elective surgery or other procedures requiring general anesthesia during the study
4. Women who are pregnant or lactating and who are planning for pregnancy during the study
5. Women who are terminally ill or inappropriate for placebo medication
6. Women with any other significant disease or disorder which, in the opinion of the investigator, may have put the patient at risk because of participation in the study, or may have influenced the result of the study, or the patient’s ability to participate in the study
7. Women with a known history of substance abuse
8. Women with a history of cannabis ingredients allergy or suspected of having an adverse reaction to cannabis
9. Women using antibiotics within the past 3 months or using NSAID drugs
10. Women consuming probiotic or prebiotic supplements

### Trial design and follow-up

#### Trial site

The prospective randomized placebo-controlled study will recruit the participants at the King Chulalongkorn Memorial Hospital (KCMH) in Bangkok, Thailand. The study is scheduled to launch approximately in June 2023.

#### Patient data confidentiality

Only the patients providing written informed consent will proceed to participate in the trial. All the data will be maintained confidentially by the research team.

#### Patient enrollment and recruitment

Patient demographic information will be reviewed by physicians using KCMH’s database. Online (e.g., on Facebook) and onsite poster (e.g., at KCMH) will be used to advertise the study. Patients will be recruited using the inclusion and exclusion criteria. The medical record database will be used to collect clinical and laboratory investigation data related to breast cancer staging, hormonal breast cancer subtypes, baseline symptoms, current medication including chemotherapy regime and other herbal use, emetic condition, history of surgery, history of other cancers, sites of metastasis, history of smoking, history of alcohol use, history of drug abuse, and history of radiotherapy.

For the recruitment, we will give the trial information to potential participants using surgeons and nurses in each surgical ward or outpatient clinic and a participant information sheet to confirm the participant’s understanding of the trial objective, procedure, benefits, and risks. The recruitment will be performed in the Thai language as native. If potential participants are willing to participate in the trial, they will be asked to sign an informed consent form with the understanding that they can quit the test at any time without any consequence.

#### Patient allocation and blinding

The recruited patients will be randomly allocated into 3 groups: Ganja Oil (THC 2 mg/mL; THC 0.08 mg/drop and CBD 0.02 mg/drop), Metta Osot (THC 81 mg/mL; THC 3 mg/drop), and placebo oil groups. The allocation will be conducted with a 1:1:1 ratio using block randomization with a block of five by the table of random numbers for creating randomization ID. The research team will provide the randomization code in an opaque envelope to conceal allocation, mask clinicians, data collectors, and patients. Following the cannabis practice patterns of the preliminary study (24), all the participants will be received cannabis or placebo oil once per day before bedtime for 90 days.

#### Visits and Follow-ups

There will be four visits for the recruited participants during the trial. At the first visit, research investigators will perform the enrollment by checking the eligibility screening, informed consent form, and allocation plan. After setting the random assignment, further assessment will be conducted using the case report forms approved by the IRB. Then, the blood, saliva, stool, and skin samples will be collected as a baseline and start the treatment a day 1 trial. The second visit will be on another day to assess the adverse drug reaction by a medical officer. The first follow-up treatment, as the third visit, will be on day 30, and the participants will be checked. Day 90 will be the last visit and second follow-up of the daily treatment at which all the participants will be assessed, and the blood, saliva, stool, and skin samples will be collected post-treatment.

After the trial, patients will be asked to stop taking cannabis oil from any sources. They will be monitored for another month for withdrawal symptoms.

#### Trial timepoints

Assessment of adverse drug effects will be collected with 4 time points: 0^th^, 1^st^, 6^th^, 12^nd^ weeks of the trial day. The pain symptoms, QoL and diet pattern will be recorded with 3 time points: 0^th^, 6^th^, 12^nd^ weeks. The blood collection (3 mL) for pharmacokinetic and pharmacogenetic tests will be performed on 2 time points: 0^th^, 6^th^ weeks. The blood (10 mL), feces, skin scrapping, and saliva samples will be taken for biochemistry tests (plasma/ serum), microbiome (gut/ skin/ saliva), cytokines (plasma) and bacterial metabolites (serum/ feces) will be analyzed at 2 time points: 0^th^, 12^nd^ weeks. The overview of the tests and visits of the clinical trial was shown on the following Table 1.

**Table 1.** SPIRIT schedule of enrolment and assessments

| # | Timepoint | Week-0<br>Baseline | Week-1<br>Follow-up | Week-6<br>Follow-up | Week-12<br>Follow-up |
| --- | --- | --- | --- | --- | --- |
|  |  | T1 | T2 | T3 | T4 |
| 1 | **Enrolment** |  |  |  |  |
|  | Eligibility screening | x |  |  |  |
|  | Informed consent | x |  |  |  |
|  | Allocation | x |  |  |  |
| 2 | **Assessment** |  |  |  |  |
|  | CRF01 Case Record Form<sup>a</sup> | x |  |  |  |
|  | CRF02 FW & ADR<sup>b</sup> | x | x | x | x |
|  | CRF03 ESAS | x |  | x | x |
|  | CRF04 EQ-5D-5L | x |  | x | x |
|  | CRF05 Food Frequency Questionnaire | x |  | x | x |
| 3 | **Blood Samples, Serum** |  |  |  |  |
|  | Blood Chemistry<sup>c</sup> | x |  |  | x |
|  | Cytokines<sup>d</sup> | x |  |  | x |
|  | Bacterial Metabolites<sup>e</sup> | x |  |  | x |
|  | Pharmacokinetics parameters | x |  | x |  |
|  | Pharmacogenetics parameters | x |  | x |  |
| 4 | **Feces Samples** |  |  |  |  |
|  | Fecal Microbiome Profile | x |  |  | x |
|  | Fecal Bile Acid Profile<sup>f</sup> | x |  |  | x |
|  | Fecal SCFAs<sup>g</sup> and BCFAs<sup>h</sup> | x |  |  | x |
| 5 | **Skin Samples** |  |  |  |  |
|  | Skin Microbiome Profile | x |  |  | x |
| 6 | **Saliva Samples** |  |  |  |  |
|  | Oral Microbiome Profile | x |  |  | x |

<sup>a</sup>Demographic and Epidemiological Factors: age, weight, height, waist circumference, hip circumference, neck circumference, alcohol consumption, smoking status, occupation, monthly income, method of health payment, and highest education.
<sup>b</sup>Follow up and adverse drug reaction: vital signs, physical examination, lab results to assess liver, kidney and heart function, pharmacokinetic record form, drug tolerance assessment form, and adverse event assessment form.
<sup>c</sup>Blood Chemistry: cholesterol, alkaline phosphatase, bilirubin, lactate dehydrogenase, gamma-glutamyl transferase
<sup>d</sup>Cytokines: interleukin-17, interferon-gamma, Interleukin-10, tumor growth factor-beta
<sup>e</sup>Bacterial Metabolites: lipopolysaccharide binding protein, sCD14 levels, equol, cadaverine.
<sup>f</sup>Fecal Bile Acid Profile: chenodeoxycholic acid, deoxycholic acid, lithocholic acid.
<sup>g</sup>SCFAs: butyrate, propionate, acetate.
<sup>h</sup>BCFAs: iso-butyric acid.

### Primary outcome measures

The pain level of each patient will be assessed using the Edmonton Symptom Assessment System (ESAS) form at the 4^th^ visit as primary outcome. There are 10 symptoms that should be assessed in the questionnaire and each scale is numbered 0-10, with 0 being asymptomatic and 10 being the most symptomatic.

### Secondary outcome measures

The ESAS at 0^th^, 6^th^ weeks visits will be taken as secondary outcome measures and will be evaluated all the symptoms.

For QoL assessment, the Thai version of the EQ-5D-5L questionnaire will be applied to evaluate at 0^th^, 6^th^, and 12^nd^ weeks. At each dimension, it has the coefficients of the level of severity utilized for calculating the utility score in accordance with the program of Health Intervention and Technology Assessment Program (HITAP).

Adverse effects including headache, dizziness, increase nausea and vomiting, dry mouth, diarrhea, muscle cramps, decreased appetite, sedation, somnolence, itching, or allergic reactions will be examined using the approved case report form. The patients will be required to evaluate the severity of the effects using 5-grade system, and the relationship to the study drugs by 4 statuses.

Regarding feces, skin scrapping, and saliva microbiome, the comparison of the microbial abundance at baseline and at the end of the trial for different arms will be performed to explore the microbial changes by the cannabis oil treatment in terms of pain assessment. The alteration of a variety of cytokines and different bacterial metabolites will be evaluated along with the microbiome test.

Plasma concentration of THC and CBD will be measured quantitatively to determine the pharmacokinetic and pharmacogenetic parameters.

### Safety

It has been confirmed the effectiveness of the cannabis products in the preliminary analysis and monitored the side effect at the trials.

### Statistical considerations

#### Sample size estimation

We target 30 patients in each arm, considering the maximum follow-up loss of 50%. Since the target population is metastatic breast cancer cases, questionnaires and biological samples such as blood, stool, and breast skin scrapping will be collected before and after the treatment, which could increase the withdrawal rate at any time during the trial.

#### Statistical analysis

The data collected from different questionnaires, including case report forms, will be prepared in Microsoft Excel, followed by data cleaning and extraction. The statistical analysis will be performed on an intention-to-treat basis using Stata/MP software version 16.0 (StataCorp 2017, College Station, TX). Descriptive statistics like pair t-tests for before and after-treatment groups will be carried out using mean with standard deviation for normally distributed continuous data and median with interquartile range for non-normal distributed continuous data. Categorical data will be presented as counts and percentages using the Chisquare test. Continuous data will be assessed for normal distribution using a histogram and Shapiro–Wilk test. One-way ANOVA with Bonferroni correction will be used for normally distributed continuous data, and the Kruskal-Wallis test will be used for non-normal distributed continuous data among 3 arms.

To identify the alpha diversity, Shannon’s diversity index is used for evenness and Chao 1 index for the richness of operational taxonomic units (OTUs). The Jaccard dissimilarity index is used to detect beta diversity, and it is statistically described by permutational multivariate analysis of variance using distance matrices (PERMANOVA) test. A statistically significant level is defined as p < 0.05. Correction for multiple analyses was not done, but they will not be included in the conclusion of the study as factual findings.

Sensitivity analysis using some subgroup to show robustness will be considered if the primary outcome is statistical significance.

## Discussion

With a rising incidence of 22,158 new cases occupying a prevalence of 22.8% amongst all female breast cancer cases in Thailand in 2020 (27, 28), more emphasis becomes on the longterm quality of life and measurement of patients’ symptoms using validated tools (29, 30). Thailand actively launched cannabis oil in the market in 2018, subsequently, the FDA (Food and Drug Administration) authorized all hospitals running under the Public Health Ministry for medical cannabis to be available on prescription to patients for approved conditions like cancer chemotherapy for relief of pain, to counter inflammation, and so on (31). Following the preliminary analysis of the Thai Cannabis PQ study, it is expected to improve the chemotherapy-related symptoms, their quality of life, and the adverse effects.

The CBC2 study is a 1-year prospective study, and it is designed as a randomized, double-blinded, placebo-controlled trial that would be conducted on current female breast cancer cases attending a tertiary medical school. This is a continual clinical trial to obtain additional information; the suggested therapy is a different drug for treating breast cancer, and it has predictable and controllable side effects.

Since some studies found a particular pattern of bacterial and fungal profiles in different breast cancer subtypes (16, 19), we expected to identify the similar microbial profile in hormone-dependent and independent breast cancer patients before and after the treatment with cannabis oil and placebo oil and evaluate the statistical changes by the treatment.

Dysbiosis is an imbalance of healthy and derogatory bacterial community structures leading to the development of chronic inflammatory conditions and cancers, particularly in the gastrointestinal tract (32, 33). Therefore, we will investigate proinflammatory cytokines such as interleukin-17 (IL-17), interferon-γ (IFN-γ), and inflammatory cytokines like interleukin-10 (IL-10), and tumor growth factor-β (TGF-β), then will explore the differences at the baseline and after the treatment.

Moreover, lipopolysaccharide-binding protein (LBP) and soluble CD14 (sCD14) levels as markers of translocation of bacteria from the gut to the blood will be measured to explore the possibility of a gut-breast cancer microbiota axis (34) and to evaluate the influence that cannabis oil treatment may have on the prevention of bloodstream transmission of oncogenic bacteria from the intestine to the breast. In addition, we will also examine levels of equol, a potent estrogenic metabolite produced by intestinal bacteria (35).

Short-chain fatty acids (SCFAs), such as acetate, propionate, and butyrate, are present in the intestine exceeding the concentration of 100 mM, and they are mainly produced by 2 major gut bacterial phyla: *Bacteroidetes* and *Firmicutes* (36). And branched chain iso-SCFAs, which play a role in membrane permeability and fluidity (37), will be investigated as bioactive bacterial metabolites in breast cancer. The SCFAs (beneficial) and iso-SCFAs (potentially harmful) will be indicators for microbial dysbiosis and bowel health (38) in participants, pre- and post-cannabis oil treatment.

Primary and secondary bile acids will be diagnosed as bioactive bacterial metabolites in breast cancer. From a range of bile acid profile, deoxycholic acid (DCA) and chenodeoxycholic acid (CDCA) at the physiological level leads to significant reductions in cell invasion, migration, adhesion, and survival but not to cytotoxicity or apoptosis (39). In addition, lithocholic acid (LCA) stimulates oxidative stress in breast cancer patients, which can reduce cancer cell proliferation (40).

The pharmacokinetic profile of cannabis oil, including THC and CBD, on routes of medication through oral administration will be examined to achieve the C_max_ of the drugs (26). As is constantly expanding the cannabis treatment, it becomes essential to predict the genes whose variations are responsible for the presence of the therapeutic and side effects of cannabis oil (41).

## Conclusions

The clinical trial will provide the efficacy of Thai cannabis oil in metastatic breast cancer female patients in Thailand, mainly in alleviating chemotherapy-related side effects. Furthermore, it will obtain secondary factors such as QoL indexes, adverse effects of the dosage, alteration of microbiome profile, bacterial metabolites, and cytokines before and after using the cannabis oil, pharmacokinetic parameters, and pharmacogenetic profiles.

## Data Availability

All data produced in the present work are contained in the manuscript.

## Data Availability

All data produced in the present work are contained in the manuscript.

## Ethics

The trial was approved by the Institutional Review Board of the Faculty of Medicine, Chulalongkorn University, Bangkok, Thailand (COA No. 1713/2022; IRB No.0548/65) on 20^th^ December 2022 in compliance with the international guidelines for human research protection as Declaration of Helsinki, The Belmont Report, CIOMS Guideline and International Conference on Harmonization in Good Clinical Practice (ICH-GCP).

## Acknowledgments

The authors are grateful to the Research Affairs, Faculty of Medicine, Chulalongkorn University, for their support.

## Author Contributions

Conceptualization: Krit Pongpirul, Mawin Vongsaisuwon, May Soe Thu, Nattiya Hirankarn Data curation: Kamonwan Banchuen, May Soe Thu, Phanupong Phutrakool, Thunnicha Ondee, Sunchai Payungporn, Pajaree Chariyavilaskul

Formal analysis: Phanupong Phutrakool, Kamonwan Banchuen, Krit Pongpirul, May Soe Thu

Methodology: Krit Pongpirul, Kamonwan Banchuen, Kulthanit Wanaratna, Pajaree Chariyavilaskul, Preecha Nootim, May Soe Thu, Sarocha Cherdchom, Sunchai Payungporn, Thunnicha Ondee

Supervision: Chanida Vinayanuwattikun, Krit Pongpirul, Mawin Vongsaisuwon, Nattiya Hirankarn, Sunchai Payungporn

Writing – original draft: May Soe Thu, Krit Pongpirul

Writing – reviewing and editing: Krit Pongpirul, May Soe Thu

